# Generalizable cursor click control using grasp-related neural transients

**DOI:** 10.1101/2020.09.03.20186973

**Authors:** Brian M Dekleva, Jeffrey M Weiss, Michael L Boninger, Jennifer L Collinger

## Abstract

Intracortical brain-computer interfaces (iBCI) have the potential to restore independence for individuals with significant motor or communication impairments. One of the most realistic avenues for clinical translation of iBCI technology is to enable control of a computer cursor—i.e. movement-related neural activity is interpreted (decoded) and used to drive cursor function. Both nonhuman primate and human studies have demonstrated high-level cursor translation control using attempted upper limb reaching movements. However, cursor click control—based on identifying attempted grasp—has only been successful in providing discrete click functionality; the ability to maintain click during translation does not yet exist. Here we present a novel decoding approach for cursor click based on identifying transient neural responses that emerge at the onset and offset of intended hand grasp. We demonstrate in a human participant, who used the BCI system independently in his home, that this transient-based approach provides high-functioning, generalized click control that can be used for both point-and-click and click-and-drag applications.

## Introduction

The loss of upper limb motor function due to injury or disease affects the ability to perform physical activities of daily living, including operating electronic devices. Intracortical brain-computer interface (iBCI) systems, which interpret motor intent signals from movement-related brain areas, may eventually be paired with dexterous robotic limbs (Carmena et al., 2003; Collinger et al., 2013; Hochberg et al., 2012; Velliste et al., 2008) or electrical stimulation of paralyzed limbs (Ajiboye et al., 2017; Ethier et al., 2012; Friedenberg et al., 2017) to return “natural” upper limb motor control. While these goals are not yet fully realizable in a clinical implementation, it is possible with current iBCI technology to provide high performance cursor control for use with computer-based applications (Nuyujukian et al., 2018; Pandarinath et al., 2017; Simeral et al., 2011; Weiss et al., 2019). Computer use provides a means of connecting to the world, and can greatly improve quality of life for those living with severe motor impairment by allowing access to web browsing, social media, electronic games, or text-based communication (Gilja et al., 2011; Huggins et al., 2011; Ryu & Shenoy, 2009; Wolpaw et al., 2002).

iBCI systems for motor control—including cursor control—interpret neural activity recorded from movement-related brain areas during attempted or imagined limb movement (Aflalo et al., 2015; Hochberg et al., 2006; Wang et al., 2009; Wolpaw et al., 2002). Commonly, cursor translation is controlled using neural activity related to attempted arm movements; for example, an attempted reach to the left is converted into leftward cursor movement (typically velocity). Similarly, cursor click is derived from neural activity during attempted hand grasp, where grasp is converted to a clicked state and the absence of grasp (neutral/relaxed posture) to an unclicked state (Bacher et al., 2015; Kim et al., 2011; Nuyujukian et al., 2018; Simeral et al., 2011). This approach provides the user with intuitive control, and can achieve performance levels suitable for real-world use in some applications (Nuyujukian et al., 2018). However, existing click decoding approaches have demonstrated only discrete click functionality, and not the ability to maintain click during translation. Such generalizable control is essential to achieve clinically-viable application of iBCI for full computer access.

The restriction of current decoding approaches to only discrete click control arises from a difficulty in identifying salient, continuous neural responses that are unique to grasp. There is some evidence that grasp-related features of neural activity are attenuated during attempted arm translation (Downey et al., 2018). Previous studies utilizing click decoding avoid this complication by using calibration routines that separate translation and grasp phases (Kim et al., 2011). This simplifies the problem of isolating activity related to grasp by avoiding translation-grasp interactions, but also limits the functionality of the decoder.

Here we present a novel approach to click decoding that identifies transient neural responses related to *transitions* in grasp state (i.e., grasp and release). This differs from previous click decoders, which instead attempt to continuously identify responses related to the grasp state itself (grasped or un-grasped). A participant with tetraplegia enrolled in an ongoing clinical trial used both types of click decoders to perform controlled tasks requiring point-and-click and click-and-drag functionality. We found that the transient-based decoding approach provided a high degree of control on both tasks, whereas the existing grasp state decoder could only achieve point-and-click control. These results advance the performance standard for iBCI click decoding, and highlight the potential importance of incorporating transient cortical responses into iBCI decoder design.

## Methods

### Participant

The participant in this study (P2) is a man with tetraplegia caused by C5 motor/C6 sensory ASIA B spinal cord injury. The participant has some residual upper arm and wrist movement, but no hand function. Approximately five years prior to data collection for the current study, two 88-channel microelectrode arrays (Blackrock Microsystems, Salt Lake City, UT) were implanted in the hand and arm areas of motor cortex. He also had two 64-channel arrays implanted in somatosensory cortex (Flesher et al., 2016), which were not used for this study. Informed consent was obtained prior to performing any study-related procedures.

### Data acquisition

All experiments were performed using an at-home portable iBCI system (Blackrock Microsystems, Weiss et al., 2019). This study was conducted under an Investigational Device Exemption from the Food and Drug Administration and approved by Institutional Review Board at the University of Pittsburgh (Pittsburgh, PA), registered at https://ClinicalTrials.gov (NCT01894802). The system uses digital Cereplex-E headstages and a portable NeuroPort Signal Processor (Blackrock Microsystems), connected to a medical-grade tablet. As reported previously, this system achieves comparable performance to typical in-lab systems. Briefly, neural signals collected by the portable system were filtered using a 4^th^ order 250 Hz high-pass filter, logged as threshold crossings (−4.5 RMS) and binned at 50 Hz. The binned counts were then convolved online with a 440ms decaying exponential filter to provide a smoothed estimate of firing rate.

Due to contact restrictions caused by the COVID-19 pandemic, the participant performed all sessions independently at his home, mostly under supervision from researchers via telecommunication. An in-house Matlab-based program (Mathworks Inc) was used to automate the session, progressing through decoder calibration and evaluation tasks. Caregivers were trained to connect the digital headstages and perform battery changes for the tablet when necessary. However, the participant performed all other elements of system operation using the tablet touchscreen, which mostly entailed running the program, selecting the task from a dropdown menu, and pressing on-screen buttons to progress through each phase of the task. Once the participant was comfortable using the system, he performed some sessions completely independently, with no assistance from researchers. All data was collected between 1795 and 1909 days post-implant.

### Dimensionality reduction

Recent work in nonhuman primate neurophysiology indicates that neural population activity in motor cortex during upper limb movement can be captured by a relatively small number of correlation patterns (Churchland et al., 2012; Degenhart et al., 2020; Gallego et al., 2017; Sadtler et al., 2014). To mitigate concerns of overfitting during decoder training and take advantage of the apparent stability of low-dimensional components (Gallego et al., 2018, 2020), we reduced the neural activity to a 20-dimensional state space using factor analysis. On each session we calculated factor weights from data collected during each initial decoder calibration routine (observation), and then used those weights to reduce incoming data to a 20-dimensional state space. The activity within this reduced state space was then used to train the click decoders.

### Decoder calibration tasks

We tested two types of calibration routines for translation and click: (1) discrete click center-out, and (2) sustained click center-out. All decoder calibration occurred at the beginning of the session and consisted of two components: observation followed by partially-assisted brain control (see *Translation decoding*). During the observation period, the participant observed the cursor as it moved under computer control between targets on the screen and transitioned between click states. The participant was asked to perform covert (i.e. imagined) movements with his right arm corresponding to the cursor translation (e.g. move arm to the left when the cursor moves to the left), and attempted grasp corresponding to cursor click (i.e. grasp for click, release for unclick). The participant’s hand is naturally in a clenched, palmar grasp posture due to hypertonia, and he reported that his motor imagery for “grasp” and “release” was best described as isometric force production (“squeeze” and “relax”) rather than finger movements.

#### Discrete click calibration task

A schematic of the task is shown in Fig 1a. Each trial (40 total) began with the cursor in the central target. One of eight outer targets then appeared, and after a short delay (0.5s) the cursor moved with a bell-shaped velocity profile to the outer target. A voice then cued the participant to “click”, followed approximately three seconds later by “release”. During the clicked period, the cursor changed from an open to filled circle. After release, the cursor returned to the center target to begin a new trial. Due to click and release occurring consecutively at the outer target, this calibration task only included cursor translation while the cursor was in the unclicked state (Fig 1a, bottom). This calibration design closely mirrors the one used previously for demonstrating point-and-click control (Kim et al., 2011)

**Figure 1.**
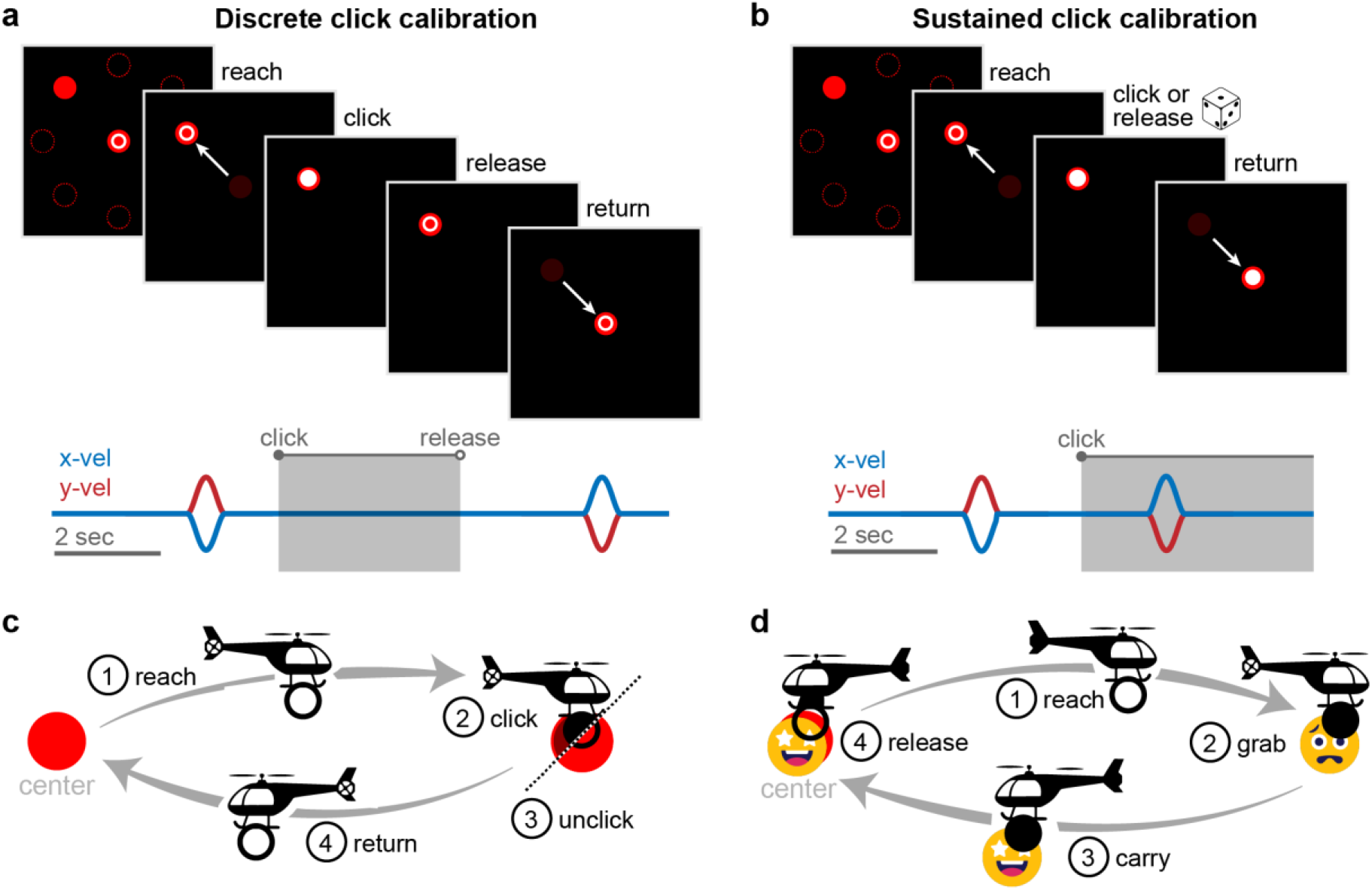
Calibration and evaluation tasks. **(a)** *Top*: Discrete click calibration task. On each trial, the cursor moved to one of eight outer targets, clicked and then unclicked (with verbal “click” and “release”) cues, and then returned to the center. *Bottom*: Example cursor velocities and click state. **(b)** *Top*: Sustained click calibration task. On each trial, the cursor moved to one of eight outer targets, then either clicked, unclicked, or remained the same before returning to the center. The transition between clicked and unclicked states was randomly selected on each trial. *Bottom*: Example cursor velocities and click state. Note that unlike the discrete click calibration task, cursor translation occurred for both clicked and unclicked states. **(c)** Point-and-click evaluation task schematic. The participant moved the cursor from the center target to the outer target (one of eight center-out target locations; rightward target in this example), clicked and released, then returned to the center. **(d)** Click-and-drag evaluation task schematic. The participant moved the cursor to the outer target (rightward target in this example), clicked to grab it, then dragged it back to the center target (both targets overlapping) before releasing.

#### Sustained click calibration task

A schematic of the task is shown in Fig 1b. The sustained click calibration was very similar to the discrete click calibration, except each trial did not contain both click and release cues. Instead, the behavior of cursor click (click or release) at the outer target was chosen at random. If the selected action was redundant (e.g. a potential “click” cue when the cursor was already in the clicked state), no cue was delivered and the trial proceeded to return to the center. Importantly, this calibration paradigm resulted in cursor translation during both clicked and unclicked states (Fig 1b, bottom).

### Decoder evalution tasks

We aimed to test the ability of the click decoders to generalize across the two main functions necessary for full click function: point-and-click (discrete click), and click-and-drag. To improve user engagement, the tasks were stylized as helicopter-based arcade games. Schematics of the two tasks are shown in Figure 1c,d. During each session, the participant first completed sixteen trials of the click-and-drag task with each decoder, followed by sixteen trials of the point-and-click task with each decoder. This sequence then repeated, resulting in thirty-two trials for each decoder/task combination.

#### Point-and-click

The participant began a trial by moving the unclicked cursor to the center target. An outer target then appeared at one of 8 radial locations and the participant moved the cursor to the target. Once the cursor entered the target, he attempted to click and immediately release without leaving the target. A trial was unsuccessful if: a click occurred before reaching the target (early click), the cursor left the target before release (drag out), or he remained in any single phase longer than twenty seconds (timeout). In the case of a drag out failure, the cursor was automatically returned to the center target. During the task, early clicks did not trigger immediate trial failure, and the participant could continue to attempt the task. However, trials with early clicks were retroactively judged as failed trials.

#### Click-and-drag

As with the point-and-click task, each trial began when the unclicked cursor entered the center target. An outer target (stylized as a “worried” face) then appeared at one of eight radial locations and the participant was tasked with clicking on the target (cursor overlapping with outer target), dragging it back to the center target (outer target overlapping with center target), and then releasing. A trial was considered unsuccessful if: a click occurred before reaching the target (early click), the dragged target was dropped before returning to the center (drop), or he remained in any single phase longer than twenty seconds (timeout). As in the point-and-click task, early clicks did not trigger immediate trial failure, but were judged as such post-hoc.

### Neural components of click (grasp)

To inform the development of our novel click decoder, we first aimed to identify prominent neural activity patterns related to click (attempted/covert grasp) observed during the discrete click calibration task. To do this, we performed an extensive grid search for unique activity patterns across all potentially relevant temporal windows encompassing grasp and release (Fig 2). For each step of the search we selected a window start and window end relative to each grasp event (100ms increments) and assigned class labels to the neural factors (class A if within window, class B otherwise). We then fit a linear discriminant analysis (LDA) classifier on the resulting dataset, thus attempting to isolate the activity observed within the selected window. Using 10-fold cross-validation, we obtained the resulting performance of the classifier, measured using Matthews Correlation Coefficient (MCC):

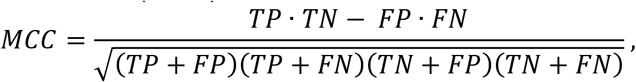

where *TP*, *TN*, *FP* and *FN* represent the true positives, true negatives, false positives, and false negatives, respectively. Due to its symmetry, the MCC metric provides a good indicator of classification performance even for highly unbalanced datasets (as is the case here for very small temporal windows). However, MCC can still display biases due to class imbalances (Zhu, 2020). To address this limitation and improve comparisons of classification performance across window sizes with different inherent class imbalances, we introduce a slightly adjusted version of MCC:

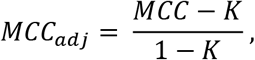

where *K* is the minimum MCC achieved across all classifications with the same class imbalance(i.e. window size).

**Figure 2.**
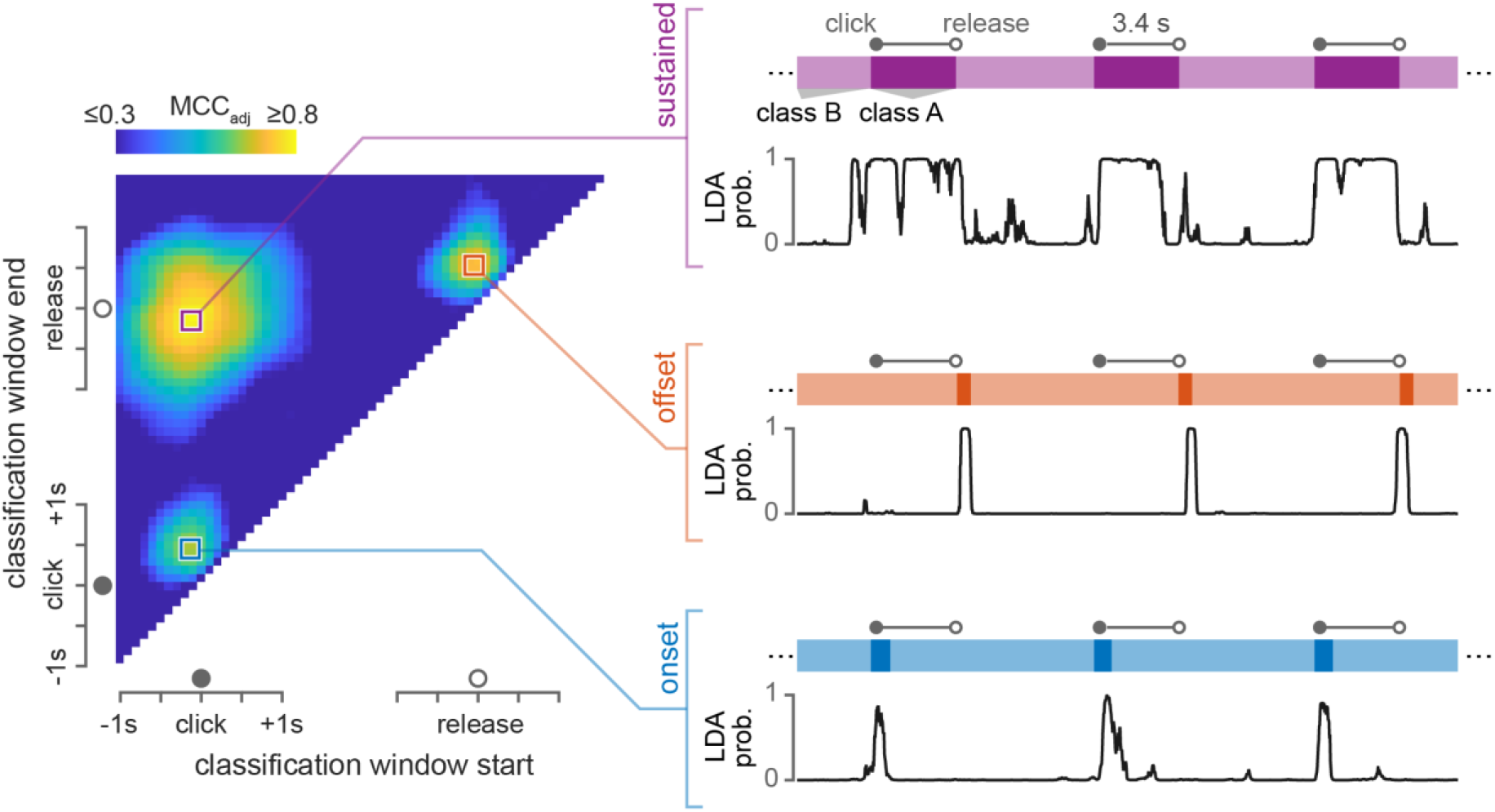
Identifying neural components related to grasp. *Left*: Discriminability of neural activity in various temporal windows around click and release. Each point represents a temporal window relative to click/release, which was used to assigned class labels to the neural data. The color at each point represents the performance (adjusted Matthews Correlation Coefficient; MCC_adj_) achieved by an LDA classifier in isolating neural activity from within the given window (10-fold cross-validated). *Right*: Example classification probability traces from the three local maxima identified through this grid search process: sustained response (purple), offset transient (orange), and onset transient (blue). Colored bars represent the target class labels for three sample trials. Black probability traces reflect the probability of class A, as output by each LDA classifier.

After sweeping across all windows from 1 second pre-click to ∼1.5 seconds post-release, we observed three local maxima—indicating three separate neural responses related to click/grasp—which are highlighted in Fig 2. The first was a window starting around the time of click onset and ending around the time of release (Fig 2, purple), indicating a neural component related to sustained grasp. The second was a window starting just before release and ending about half a second after release (Fig 2, orange), and the third was a window starting just before click and ending about half a second after click (Fig 3, blue). These two components represent transient responses at the offset and onset of grasp, respectively. Note that absolute MCC_adj_ score does not necessarily indicate the magnitude or salience of the associated neural response. Small trial-by-trial variation in the timing of an attempted action (and the corresponding neural response) will have a significantly greater impact on classification performance for short time windows (e.g. Fig 3, orange or blue) than for long windows (Fig 3, purple). However, this sensitivity to temporal variability is only present during offline classifier training, which assumes fixed relationships between the cues and the neural responses, and does not predict the performance during asynchronous online control.

### Sustained click decoder

The sustained neural response that occurs during grasp (identified in Fig 2) has been used in previous approaches to click decoding, in which a classifier is used to directly output the click state (Kim et al., 2011; Simeral et al., 2011). We replicated this approach to provide a baseline comparison of existing click decoder functionality. To calibrate the decoder, we used the click state (clicked/unclicked) at each point in the calibration task to assign class labels and then trained a simple LDA classifier on the 20-dimensional neural state (factors). During online control, we applied the classifier at each time point (20ms) and directly mapped the output to the click state.

### Transient-based click decoder

While previous approaches to click decoding utilized sustained neural responses during grasp, we aimed to instead use the transient responses observed at the onset and offset of grasp (Fig 2). To do this, we trained two independent classifiers, which then ran concurrently during online control to update changes in click state. To train each transient classifier, we first identified the optimal time window for a given session. From the results in Fig 2, we found that the transient responses (observed in the smoothed estimate of neural firing rates, see *Data acquisition)* were only about half a second long. These short time windows meant that idiosyncrasies in the user’s approach during calibration—which might vary across days or across subjects—could significantly impact the classification. For example, a user might attempt to grasp immediately upon hearing the audio cue to “click”, or might wait until visual feedback of the cursor changing from the unclicked to clicked state. For this reason, on each session we trained the transient classifiers using a limited grid-search approach similar to the one outlined in the section *Neural components of click (grasp)*. However, rather than search the entire parameter space, we restricted the search to smaller time periods. For both grasp and release, we swept through a range of window centers (−1.0s to +1.0s relative to onset/offset, 0.1s increment) and window widths (0.2s to 2s, 0.1s increments). For each window, we computed the output probabilities from the resulting LDA classifier (10-fold cross-validation) and calculated the MCC for probability thresholds between 0.1 and 0.9 (increments of 0.1). We then selected the time window with the greatest cumulative MCC (summing across all thresholds).

During online control, we applied a simple heuristic to convert the transient classifier outputs to click function. If the cursor was unclicked and the grasp onset transient probability exceeded both 0.2 and the release transient probability, the cursor entered the clicked state. If the cursor was in the clicked state and the release transient probability exceeded both 0.2 and the grasp onset transient probability, the cursor entered the unclicked state.

### Translation decoding

The focus of this study was decoding click, rather than cursor translation. To maintain consistent translation performance in combination with both tested click decoders, we used an optimal linear estimator (OLE) approach to decode cursor velocity. We have previously used this kinematic decoding approach to demonstrate high-quality control for two-dimensional cursor movement (Weiss et al., 2019) and up to ten-dimensional arm/hand movement (Collinger et al., 2013; Wodlinger et al., 2014).

Following our previous approaches for kinematic decoding, we followed up each of the calibration routines (discrete click or sustained click) with a set of partially-assisted online brain control trials. These trials (40 total) followed the same center-out format, but with movement velocity controlled by the OLE translation decoder (restricted to the target axis). The data collected during this assisted set was used to fit a new OLE translation decoder, but was not used for any aspect of click decoder calibration.

## Results

Participant P2 was able to achieve sufficient translation control under all conditions to make accurate cursor movements to the targets (Fig 3). This allowed us to use overall task performance as a gauge of relative click functionality between the two tested decoders.

**Figure 3.**
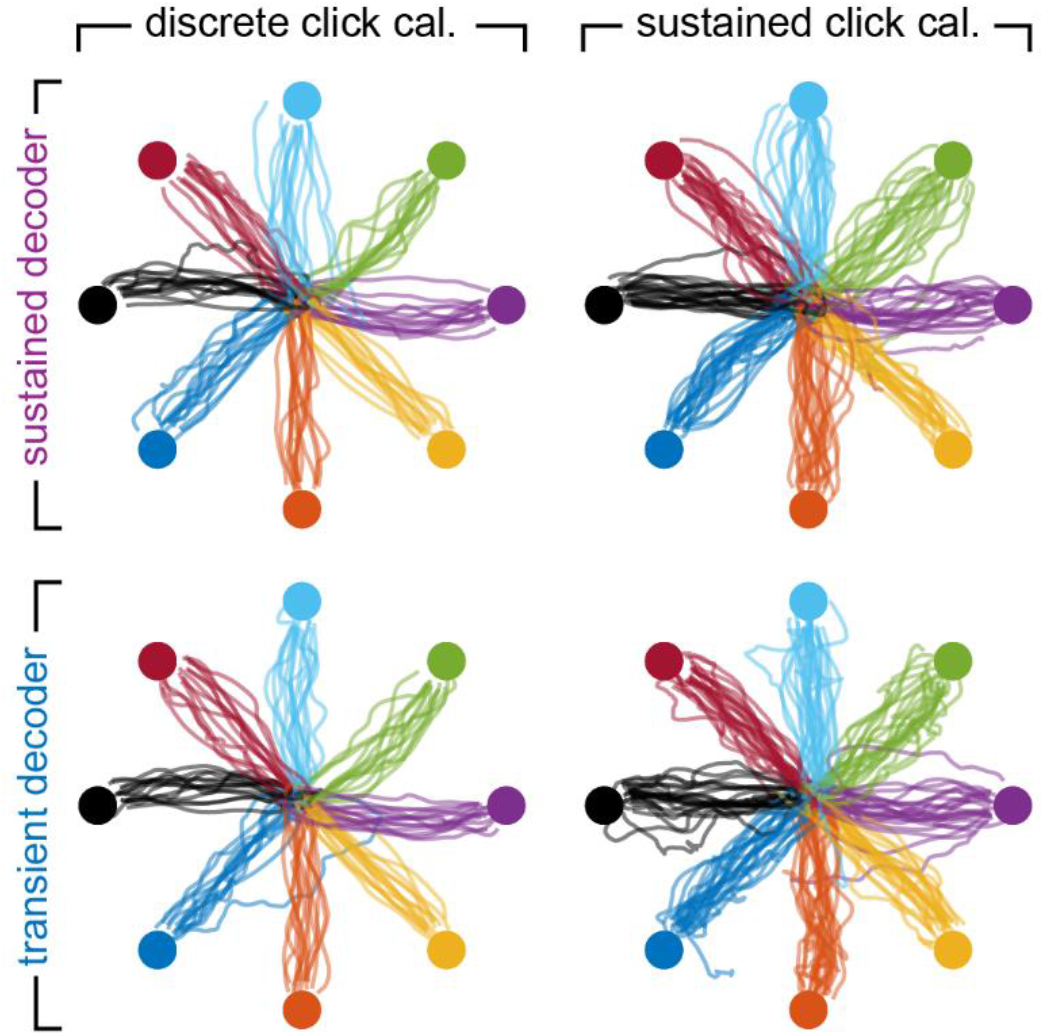
Translation control. Average cursor trajectories (each line comprises four trials) during the reach phase, separated by calibration routine (discrete or sustained click) and decoder (sustained or transient). Each subplot contains trajectories from all sessions, including both the point-and-click and click-anddrag tasks.

### Point-and-click task performance

During the point-and-click task, the sustained click decoder was only effective when trained on the discrete click calibration task (Fig 4a, left). This condition is equivalent to previous demonstrations of point-and-click control, including the same training paradigm, decoding approach, and evaluation task (Kim et al., 2011). Almost all trials of this type fell into two categories—success and early click—with roughly equal probability. The relatively low success rate in comparison to previous studies is likely because we only examined the first click on each trial when determining success or failure rather than allowing multiple attempts following an errant first click. When trained on the sustained click calibration task, the sustained decoder failed to provide adequate control, with a high occurrence of early clicks. This indicates that sustained grasp-related neural activity is not easily isolated from translation-related signals during simultaneous control, which matches results from a previous study from our group (Downey et al., 2018).

**Figure 4.**
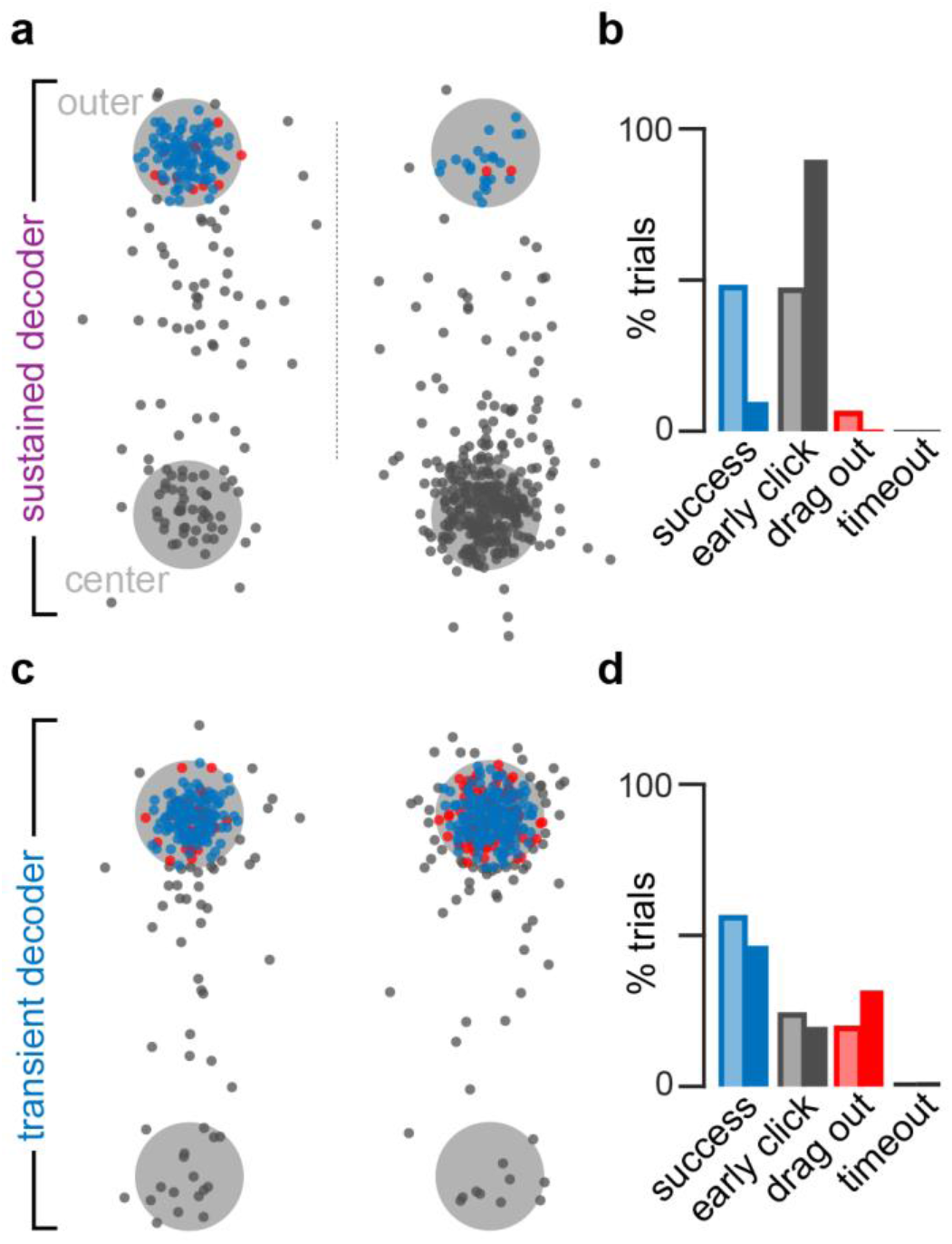
Point-and-click performance. **(a)** Click locations (rotated to align across target directions) during sustained decoder trials trained using discrete click calibration (left) or sustained click calibration (right). Blue points represent click locations on successful trials. Black points represent the click locations on trials with initial clicks outside of the target. Red points represent failed trials in which the click occurred inside the target, but the cursor left the target before release. **(b)** Histogram of outcomes for sustained decoder trials following discrete click (light) and sustained click (dark) calibration. **(c)** Same as in (a) for the transient-based decoder. **(d)** Same as in (b) for the transient-based decoder.

Unlike the sustained click decoder, the transient-based click decoder was successful at providing point-and-click functionality regardless of calibration routine (Fig 4c,d). It did display a higher incidence of “drag out” failures(i.e. failing to unclick before leaving the target). However, this can likely be attributed more to limitations in translation control than to a failure of click control. From the click locations shown in Fig 4c, the “drag out” failures almost exclusively occurred on trials where the participant clicked on the outer edge of the target. Thus, those trials appear to reflect a failure in stabilizing the cursor, and not necessarily a failure in release control.

### Click-and-drag task performance

The sustained click decoder was unable to provide any meaningful drag functionality, regardless of calibration routine (Fig 5a,b). However, the types of failures depended on the calibration routine. When trained on discrete click calibration, the participant was able to successfully reach the outer target on a significant number of trials. However, he was unable to maintain click during translation back to the center target, and almost every trial ended with an early drop. This failure to maintain the click is a result of the limitations caused by the calibration routine. The discrete click calibration routine only included interleaved translation and click. Thus, the resulting classifier was not able to generalize to the condition in which translation coincided with click. When trained on the sustained click calibration routine, the sustained click decoder behaved equally poorly, but the failures almost entirely resulted from early click—this failure type corresponds to false positives during grasp classification.

**Figure 5.**
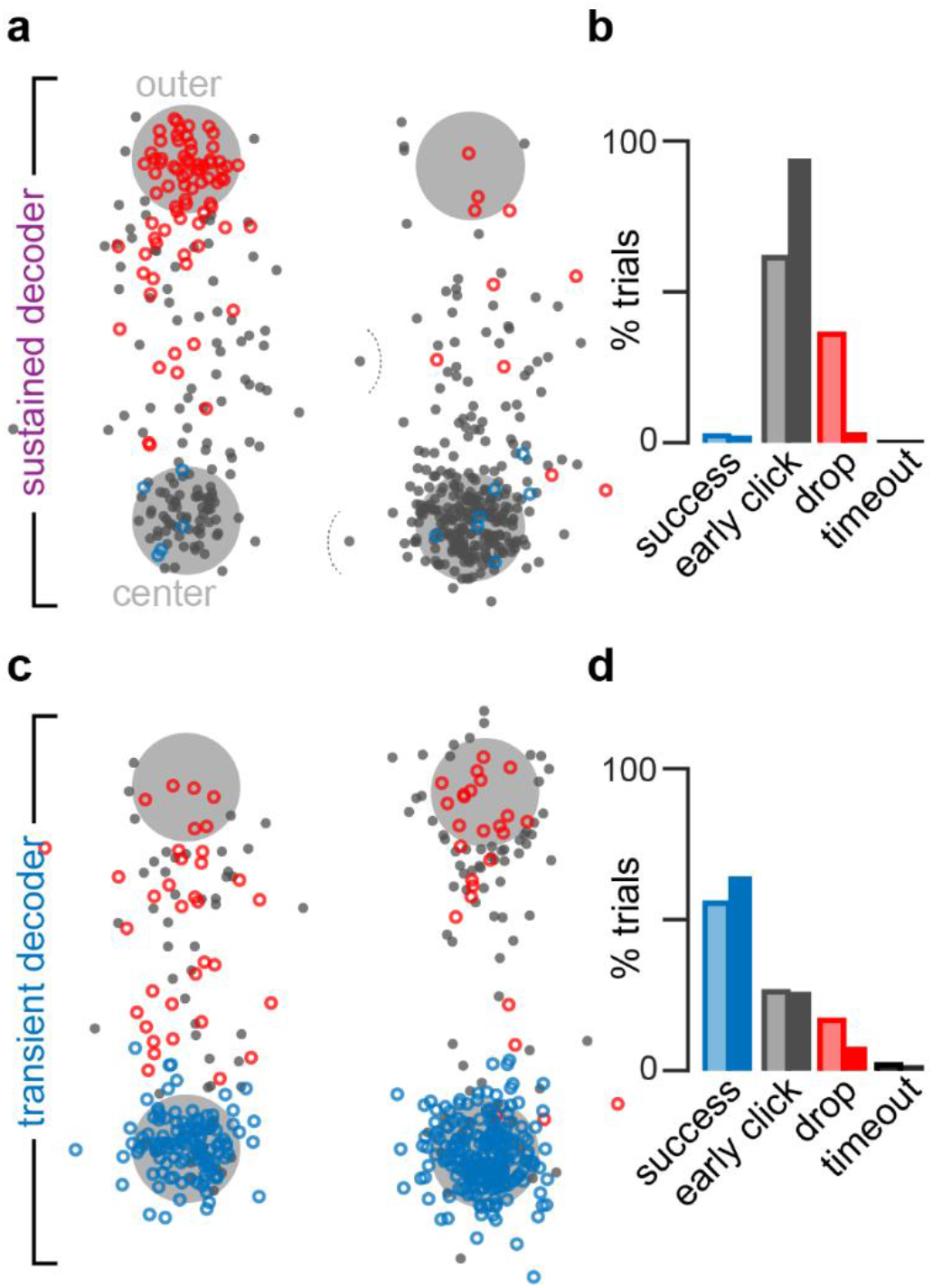
Click-and-drag performance. **(a)** Key click and release locations (rotated to align across target directions) during sustained decoder trials trained using discrete click calibration (left) or sustained click calibration (right). Open blue points represent release locations on successful trials, where the participant successfully dragged the outer target back to the center. Black points represent the click locations on failed trials with initial clicks outside of the target. Red points represent release locations on failed trials in which the participant grabbed the outer target, but released before reaching the center target. **(b)** Histogram of outcomes for sustained decoder trials following discrete click (light) and sustained click (dark) calibration. **(c)** Same as in (a) for the transient-based decoder. **(d)** Same as in (b) for the transient-based decoder.

The transient-based decoder provided high click-and-drag functionality regardless of calibration routine (Fig 5c,d). As during the point-and-click task, the most common failure was an early click before reaching the target. However, especially for the decoder trained following the sustained click calibration routine (Fig 5c, right), the majority of early clicks occurred just outside of the outer target, and thus appear to reflect inadequacies in translation control rather than click control. The decoder trained from sustained click calibration also had a lower incidence of drops (17% vs 8%, Fig 5d), suggesting slightly better release control. Together, these results indicate that while the performance of a transient-based decoding approach is largely invariant to the calibration task, a calibration routine involving sustained click (i.e. translation in both clicked and unclicked states) may lower the incidence of both unintentional clicks (Figs 4d, 5d) and unintentional releases (Figs 5d).

### Additional control metrics

The results in Figs. 4 and 5 reflect the strictest possible success criteria. As described in *Decoder evaluation tasks*, early click errors during task performance did not actually trigger trial failure. To evaluate performance in a more forgiving framework, we recalculated the overall performance metrics after allowing for multiple clicks (Fig S1), which is equivalent to the participant’s online experience while performing the tasks. For the transient decoder, this resulted in an increase in point-and-click success rates to 72% (discrete click calibration) and 57% (sustained click calibration) and click-and-drag success rates to 72% and 82%. For the sustained decoder, point-and-click success rates increased to 91% and 90%, but click-and-drag rates increased only to 5% and 16%. The most striking change in performance was the improvement in point-and-click performance by the sustained decoder (from 10% to 90%). However, the participant performed many unnecessary clicks in this condition, with 46% of successful trials containing at least five clicks (Fig S2). Trials with the transient decoder (trained using either calibration routine) contained only one or two clicks on > 95% of trials, indicating that even though the success rate was lower than the sustained decoder, it provided more reliable and consistent control.

In addition to total successes, we also investigated the temporal aspect of control achieved by each decoder. To summarize general performance speed, we calculated the target acquisition rate (number of successful trials divided by the total task time, excluding intertrial periods) for each sixteen-trial block of the point-and-click and click-and-drag tasks (Fig 6a). Acquisition rates varied considerably even within condition, which reflects cross-session variability in both click control and translation control. The sustained click decoder only achieved consistent, meaningful control on the point-and-click task, and only when trained using discrete click calibration (median rate of 3.6 successes/minute). This rate was not significantly different from the rate achieved by the transient decoder trained using discrete click (p = 0.63, Mann-Whitney U-test) or sustained click (p = 0.66, Mann-Whitney U-test) calibration. The variance in performance was also not significantly different (discrete click: p = 0.29, sustained click: p = 0.08, F-test)

For the point-and-click task, the transient click decoder achieved a median acquisition rate of 3.1 successes/minute when trained using discrete click calibration, and 3.0 successes/minute when trained using sustained click calibration. Thus, the calibration paradigm had no effect on median acquisition rate (p = 0.76, Mann-Whitney U-test). However, performance on this task was more consistent across sessions when trained using discrete click calibration (σ = 1.2 compared to σ = 2.7, p = 0.006, F-test; open vs closed circles in Fig 6a). For the click-and-drag task, the transient click decoder achieved a median acquisition rate of 3.7 successes/minute when trained using discrete click calibration and 4.4 successes/minute when trained using sustained click calibration. These rates were not significantly different from each other in median (p = 0.21, Mann-Whitney U-test) or cross-session variance (p = 0.87 F-test). These results indicate that the transient decoder, unlike the sustained decoder, provided generalizable control, and allowed the participant to achieve consistent performance across tasks.

**Figure 6.**
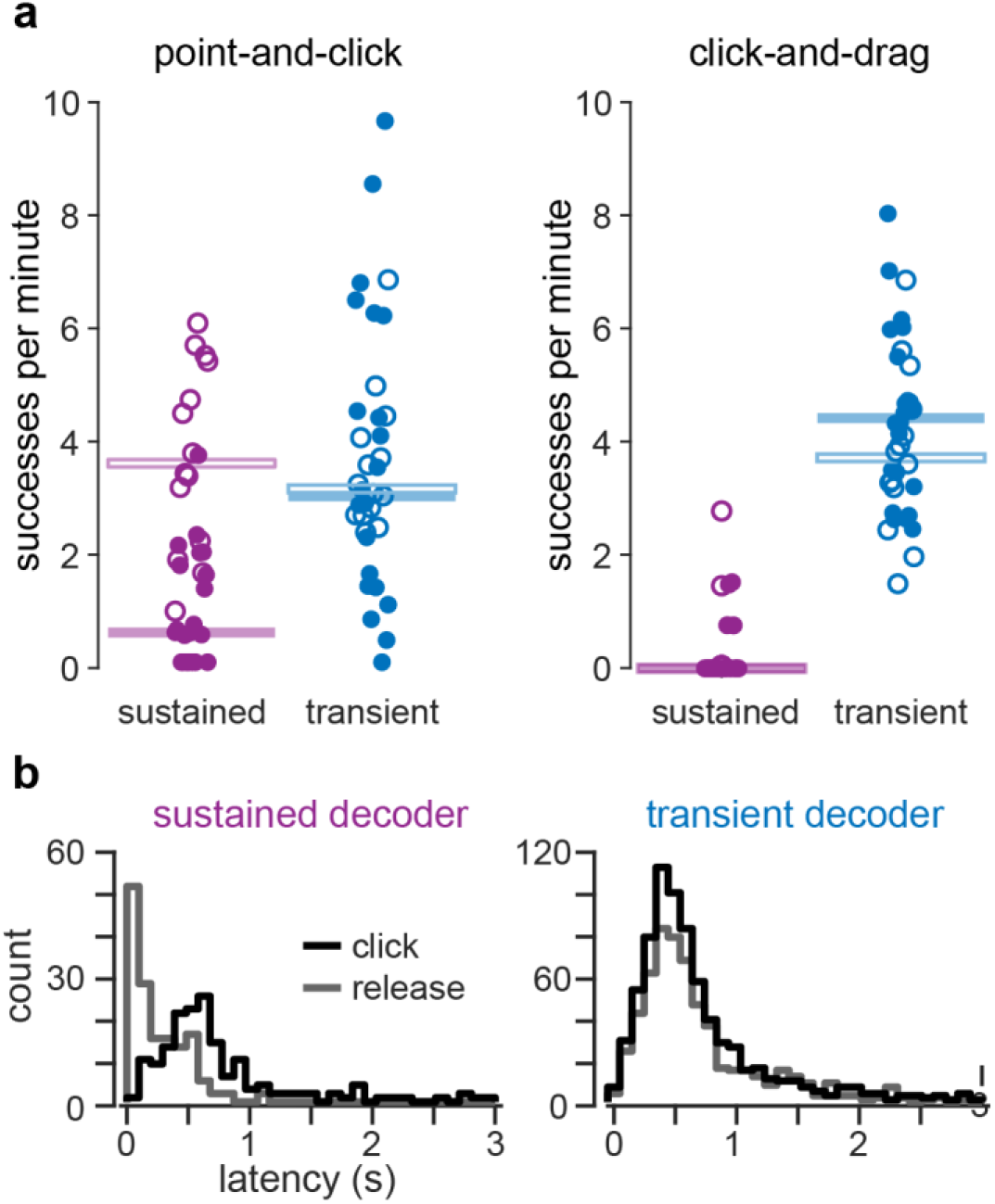
Control timing **(a)** Success rates achieved during the point-and-click and click-and-drag tasks for the sustained and transient decoders. Open circles correspond to discrete click calibration sessions and closed circles to sustained click calibration sessions. Each point represents a sixteen-trial block (two per session). Horizontal bars denote the median success rate of each group **(b)** Latency of click and release commands on successful trials from both tasks. The latency was calculated as the time delay between when a click/unclick event was possible (e.g. entering the outer target) and when it occurred.

To better analyze the behavior of each click decoder during control, we calculated the latency of click and release responses across all successful trials on both tasks (Fig 6b). On each successful trial we identified the time lag between when click or release was possible, and when it actually occurred. For both tasks, click latency thus represents the time between when the cursor entered the outer target and when click occurred. Release latency during the point-and-click task is simply the time between click and release, and for the click-and-drag task it is the time between completion of the drag (outer target coinciding with center target) and the release. The average click latencies were not different between the two decoders (p = 0.88 Mann-Whitney U-test), with a median click latency of 0.58 seconds for the sustained decoder and 0.56 seconds for the transient decoder. However, the release latencies differed significantly (p < 10^−40^, Mann-Whitney U-test), with a median release latency of 0.12 seconds for the sustained decoder and 0.56 seconds for the transient decoder. Note that for the transient decoder, the release latencies mirrored the click latencies (p = 0.61, Mann-Whitney U-test), indicating that both resulted from similar volitional control. Sustained decoder release latencies were skewed significantly lower than the click latencies (p < 10^−28^, Mann-Whitney U-test). The distribution of release latencies comprised almost entirely point-and-click trials (see breakdown of successful trials, Fig 4a,b), and indicates that for point-and-click function, a sustained click decoding approach can provide shorter click durations and improve the speed of button clicking. However, this advantage comes at the cost of generalizability, as the click cannot be maintained during cursor translation.

### Salience of grasp-related neural responses underlying click control

The behavioral results from the point-and-click and click-and-drag tasks indicate that a transient-based approach to click decoding can provide more generalizable control of cursor click. Here we examined the neural response features used by each decoding approach to better understand the cortical control of grasp and its application to decoding.

On each session, we aligned neural responses (during calibration) to click events and projected the neural activity onto the three LDA axes (see Fig 2) used by the decoders (Fig 7a,b). The resulting traces thus correspond to the main cortical responses observed during attempted grasp (see *Methods)*. Across all sessions and calibration routines (discrete click: Fig 7a, sustained click: Fig 7b), we found consistent and reliable responses related to all three grasp-related responses: onset (blue), offset (orange) and sustained (purple) grasp. However, the salience (magnitude) of the responses was not equal, with the sustained component consistently weaker than either transient response.

To compare response magnitudes across the three identified neural axes, we calculated on each trial the maximum deviation along each axis. Specifically, we found the range (difference between the maximum and minimum) observed on each trial after projecting along the onset, offset, and sustained axes (Fig 7c,d). We found that both transient responses were consistently stronger than the sustained response. The maximum deviations along the grasp onset axis were greater than the maximum deviations along the sustained axis during discrete click calibration (median onset = 3.6 a.u. median sustained = 3.1; p < 10^−38^, paired t-test) and sustained click calibration (median onset = 3.9 a.u. median sustained = 2.8; p < 10^−28^, paired t-test). The maximum deviations along the grasp offset axis were also greater than the maximum deviations along the sustained axis for discrete click (median offset = 4.1 a.u. median sustained = 3.1; p < 10^−59^, paired t-test) and sustained click (median offset = 4.1 a.u. median sustained = 2.6; p < 10^−29^, paired t-test) calibration. This result highlights the significance of transient cortical responses during grasp control, responses that were previously excluded from grasp-driven click decoding approaches.

**Figure 7.**
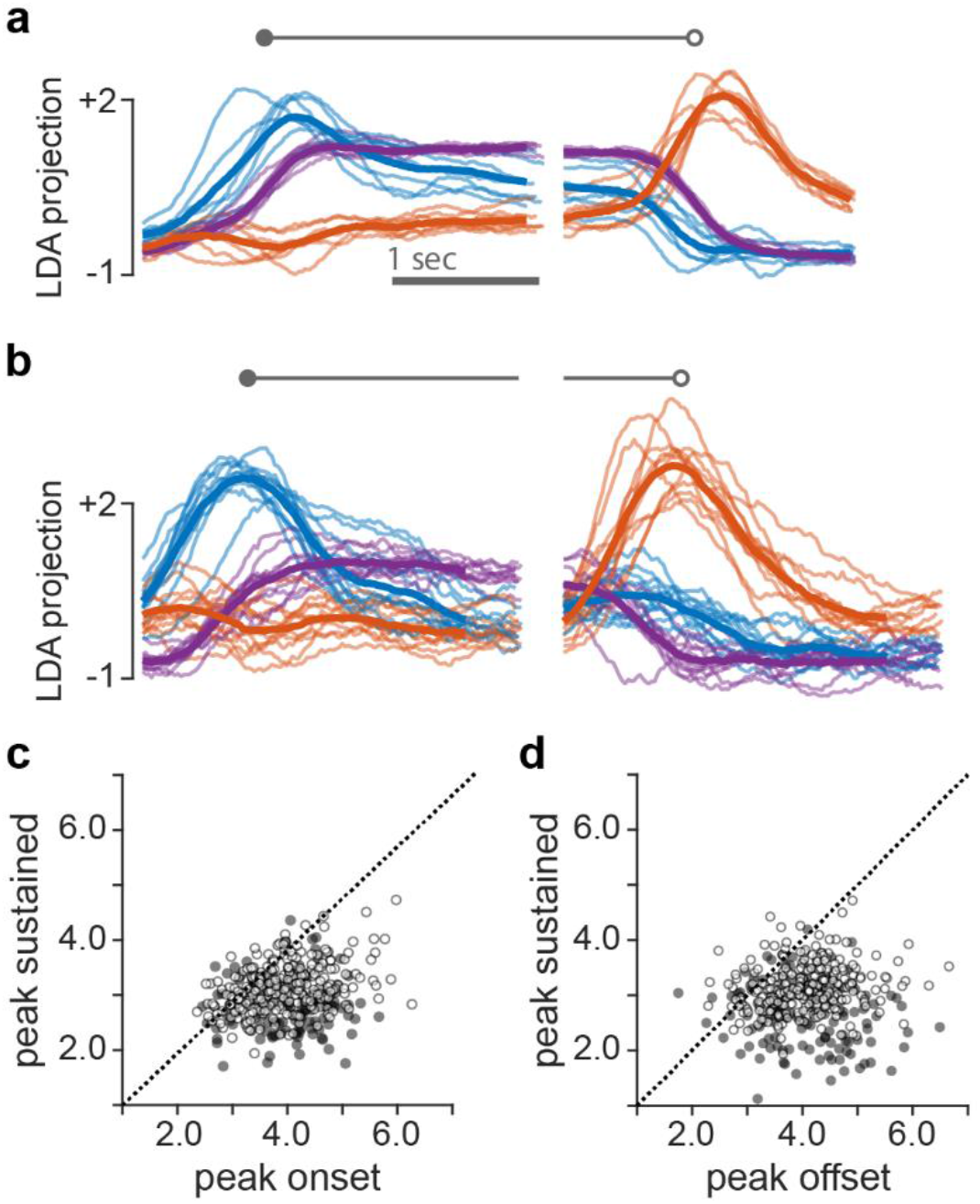
Salience of neural responses across key grasp-related dimensions **(a)** Projection of neural activity during discrete click calibration onto onset (blue), offset (orange), and sustained (purple) LDA axes. Light traces represent individual sessions. Dark traces represent cross-session averages. **(b)** Same as (a) for sustained click calibration sessions. **(c)** Comparison of peak excursions along onset and sustained axes. Open circles correspond to discrete click calibration trials, closed circles to sustained click calibration trials. **(d)** Same as (c), but comparing peak excursions along offset and sustained axes. q

### User-initiated application for mouse emulation

The participant was encouraged throughout the experimental period to use the system for applications outside of the experimental tasks outlined in this study. After performing a calibration routine (as outlined in the Methods), he was free to select a decoder and use it as a mouse emulator for any application of his choosing. Over the course of the data collection period, he used the system for this purpose fifteen times. On every occasion he chose to use the transient-based click decoder over the sustained click decoder. One common application he performed with the decoder was digital painting, which requires the ability to perform discrete clicks (for selecting brushes, colors, etc.) and also the ability to click-and-drag (for drawing lines, etc.). An example painting showcasing this control is shown in Fig 8. In addition to painting, he also used the BCI system for playing card-based computer games that require the ability to click-and-drag.

## Discussion

This study demonstrates that transient neural responses at the onset and offset of (attempted) hand grasp can be used to provide generalizable click control for intracortical brain-computer interfaces. Previous implementations of click decoding have relied on sustained cortical responses during grasp. While this approach can provide adequate point-and-click control if calibrated appropriately, it is unable to provide continuous, sustained click control during cursor translation. The transient-based approach that we introduce here provides both discrete (point-and-click) and sustained (click-and-drag) functionality, and is robust across calibration tasks.

The results presented here are from a single participant. However, transient responses appear to be ubiquitous features of cortical motor control—even during sustained isometric force production (Intveld et al., 2018; Sergio & Kalaska, 1998; Shalit et al., 2012; Smith et al., 1975). A close examination of activity patterns observed during imagined or attempted hand grasp in other intracortical human studies reveals similar onset and offset transient spikes (Rastogi et al., 2020). Thus, we believe that the transient-based decoder architecture presented in this study is taking advantage of fundamental cortical response properties and will generalize to any iBCI application with well-modulated grasp-related neural activity.

The improvement in performance achieved by the transient click decoder can be attributed to two main findings. First, the transient responses at the onset and offset appear consistently more salient (higher magnitude) than the response corresponding to sustained grasp. The larger magnitude of the responses leads to improved classification and reduces the incidence of both false positives (unintended click or release) and false negatives (poor responsiveness). Importantly, these transient responses appear to be: (1) inherent to grasp onset and offset, and (2) unique from each other. The participant reported that his attempted actions during click and unclick were squeeze (hand already in clenched posture) and relax, respectively. Thus, the observed transients do not appear to be part of a control modality separate from the sustained component (e.g. kinematics of hand closing/opening). Rather, the three response types (onset transient, offset transient, sustained grasp) seem to reflect three components of a singular, dynamic neural process underlying grasp. The lack of confusion between the two transients during classification also indicates that they reflect two unique components, rather than a single, broad response such as a global increase in firing rates.

**Figure 8.**
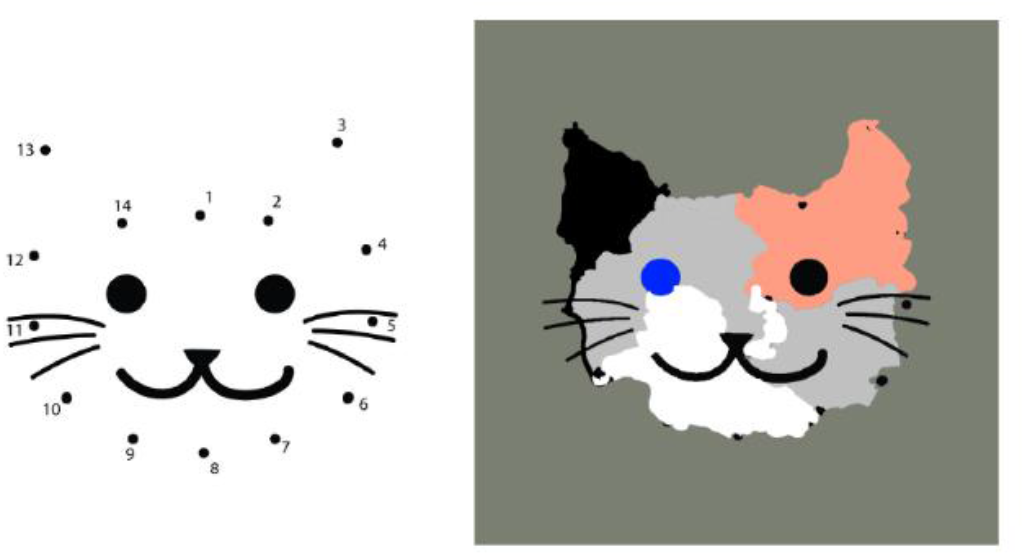
Digital painting by participant. Working from a connect-the-dots pattern (left), the participant used the transient decoder to select and apply paint colors (point-and-click functionality) and to draw outlines, erase numbering, etc. (click-and-drag functionality).

Second, by controlling click state transitions rather than the click state itself, the transient-based approach largely avoids the problem of disentangling grasp-related activity from translation during simultaneous control. Responses in motor cortex appear to modulate with a very broad range of actions, regardless of the specific recording location. During attempted click-and-drag, the neural activity thus contains components related to both cursor translation and sustained click, which complicates classification of only grasp-related activity. However, the cursor is generally at rest at the moment of click (and release), which means that classification of click state change is generally equivalent across tasks, regardless of the complexity or multimodality of control before or after. Further studies are necessary to determine whether the transient decoder can also allow for click and release in the middle of active cursor translation.

The control of click state transitions also allows for greater flexibility in modifying the control during use. Since click and release arise from separate classification processes, the thresholds can be modulated independently. For sustained click classification approaches, decreasing the threshold to make clicking easier necessarily makes releasing more difficult (and vice versa). However, for the transient click decoder, the thresholds for onset and offset can be adjusted independently as necessary to improve responsiveness (lower threshold), or prevent unintentional click state changes (higher thresholds) for both click and release. Additionally, onset and offset detection can be integrated into translation control to improve usability. For instance, leaving the target before release (drag out) was one of the most common failure modes for the point-and-click task. To prevent this type of error, translation control can be disabled when either transient click-related response (onset or offset) is detected. This immobilizes the cursor during click transitions and reduces the effect poor cursor stabilization, a common problem in iBCI (Sachs et al., 2015). Thus, a transient-based click decoding approach not only provides more generalizable control of click, but also allows for more customization to meet the needs and wants of the end user.

## Conclusion

We have demonstrated that cortical transients at the onset and offset of attempted grasp can be used to provide high-quality, generalizable click control for iBCI computer cursor applications. A participant was able to use this transient-based click decoder to achieve both point-and-click and click-and-drag functionality, which was not possible with previous click decoding approaches. The success of this transient-based approach highlights the importance of understanding the full range of response characteristics in motor cortex when developing decoding algorithms for iBCI systems. Future studies will focus on extending the dimensionality of click control (e.g. multiple button clicks) and translating the decoding approach to the control of robotic limbs to improve real-world grasp functionality.

## Data Availability

Data can be made available upon reasonable request.

## Acknowledgments

Research reported in this publication was supported by the National Institute Of Neurological Disorders And Stroke of the National Institutes of Health under Award Numbers UH3NS107714 and U01NS108922, Defense Advanced Research Projects Agency (DARPA) and Space and Naval Warfare Systems Center Pacific (SSC Pacific) under Contract N66001–16-C4051, and the UPMC Rehabilitation Institute. The content is solely the responsibility of the authors and does not necessarily represent the official views of the National Institutes of Health, DARPA, or SSC Pacific.

